# Smartphone keyboard dynamics predict affect in suicidal ideation

**DOI:** 10.1101/2023.11.29.23299169

**Authors:** Loran Knol, Anisha Nagpal, Imogen E. Leaning, Elena Idda, Faraz Hussain, Emma Ning, Tory A. Eisenlohr-Moul, Christian F. Beckmann, Andre F. Marquand, Alex Leow

## Abstract

While digital phenotyping provides opportunities for unobtrusive, real-time mental health assessments, the integration of its modalities is not trivial due to high dimensionalities and discrepancies in sampling frequencies. We provide an integrated pipeline that solves these issues by transforming all modalities to the same time unit, applying temporal independent component analysis (ICA) to high-dimensional modalities, and fusing the modalities with linear mixed-effects models. We applied our approach to integrate high-quality, daily self-report data with BiAffect keyboard dynamics derived from a clinical suicidality sample of mental health outpatients. Applying the ICA to the self-report data (104 participants, 5712 days of data) revealed components related to well-being, anhedonia, and irritability and social dysfunction. Mixed-effects models (55 participants, 1794 days) showed that less phone movement while typing was associated with more anhedonia (β = -0.12, p = 0.00030). We consider this method to be widely applicable to dense, longitudinal digital phenotyping data.

## Introduction

Traditionally, mental health assessments are administered by professionals in the clinic and therefore occur infrequently, outside the context of an individual’s daily life. The ubiquity of smartphones presents many opportunities for more frequent mental health assessments outside of the clinic.^1^ A popular and direct measure of mental state administered through smartphones are self-report questionnaire-style prompts, like ecological momentary assessment (EMA), which repeatedly sample behaviour and experiences in their natural environment, in real-time.^2^ As with any measurements that rely on active user engagement, however, EMA imposes burden on the participant and is therefore prone to attrition.^3^ Therefore, the development of passive, unobtrusive smartphone measures that predict mental state is receiving increasing attention.^4^

Quantifying behavioural phenotypes using data collected unobtrusively from wearable digital devices is referred to as digital phenotyping.^5^ One example of this approach is the open-science iOS app BiAffect.^6^ Developed by our team, BiAffect replaces the user’s iPhone keyboard. It collects keyboard typing metadata (e.g., typing speed) as well as accelerometery data (movement and orientation while typing). Previous work has shown that typing speed predicts cognitive processing speed and shows an age-modulated, diurnal pattern.^7,8^ In addition, several measures such as accelerometer displacement and autocorrect rate have been shown to predict depression or mania ratings.^6,8,9^ These findings highlight the potential of passively collected typing data in clinical contexts.

While providing unique opportunities, the inception of this new technology requires analytical workflows that can extract meaningful behavioural phenotypes from the underlying timeseries. This poses several analytical challenges. Most importantly, it is often necessary to integrate data modalities that are acquired at different sampling frequencies. This is important, for example, to validate the predictive power of digital phenotyping measures for mental health and cognition, most commonly against self-report measures.^4^ However, self-report prompts tend to occur, at most, several times a day, which forms a data stream that is very sparse compared to the hundreds of daily samples collected by smartphones. Therefore, any study that aims to validate digital phenotyping measures of mental health must first address temporal misalignment. An additional problem arises when data are high-dimensional, making dimensionality reduction techniques desirable.

The solution we employ involves: 1) applying temporal independent component analysis (ICA) to the high-dimensional modalities, 2) transforming all modalities to the same time unit through resampling or aggregation, and 3) then fusing the modalities through linear mixed-effects models as in prior work.^10–14^ Temporal ICA decomposes a multivariate time series into a limited set of components by maximising their statistical independence in the time domain.^11,12^ Crucially, ICA does not collapse the time domain, allowing classical resampling and aggregation techniques to align the generated independent components with the other digital phenotyping modalities. Additionally, ICA can compress data into a smaller number of independent components, making it ideally suited for dimensionality reduction. This means that fewer mixed-effects models need to be constructed, leading to a more parsimonious system of models that suffers less from multiple comparison corrections.

To demonstrate the value of our approach, we apply it to integrate high-quality self-report data with digital phenotyping data from the CLEAR-3 trial, a randomised controlled crossover trial that investigated how a hormonal intervention impacts menstrual cycle exacerbation of suicidal ideation and affective symptoms. The trial featured a unique clinical sample of mental health outpatients who were assigned female at birth (AFAB) and reported suicidal ideation in the past month. Participants self-reported on a large array of questionnaire items pertaining to affective, cognitive, and behavioural functioning on a daily basis and received substantial monetary compensation for the completion of daily ratings to ensure a high response rate that is not feasible in real-world applications. Meanwhile, they were encouraged to use the BiAffect iOS keyboard for the duration of the study.

We applied temporal ICA to the self-report data to distil the large number of items into fewer dimensions and predict their time course from BiAffect-derived data streams. Before running the ICA, we concatenated the self-report data of all participants along the temporal domain, both to increase the number of time steps fed into the analysis and to get a common set of independent components that applies to all participants.^12,15^ Temporal ICA then takes such a matrix of time series and decomposes it into a time-free mixing matrix and a set of components that are independent in the temporal domain. The mixing matrix specifies how the independent components combine to generate the original measures. We consequently constructed a separate data fusion model for each component, employing strict multiple comparison corrections. An overview of our approach is given in Figure 1.

**Figure 1:**
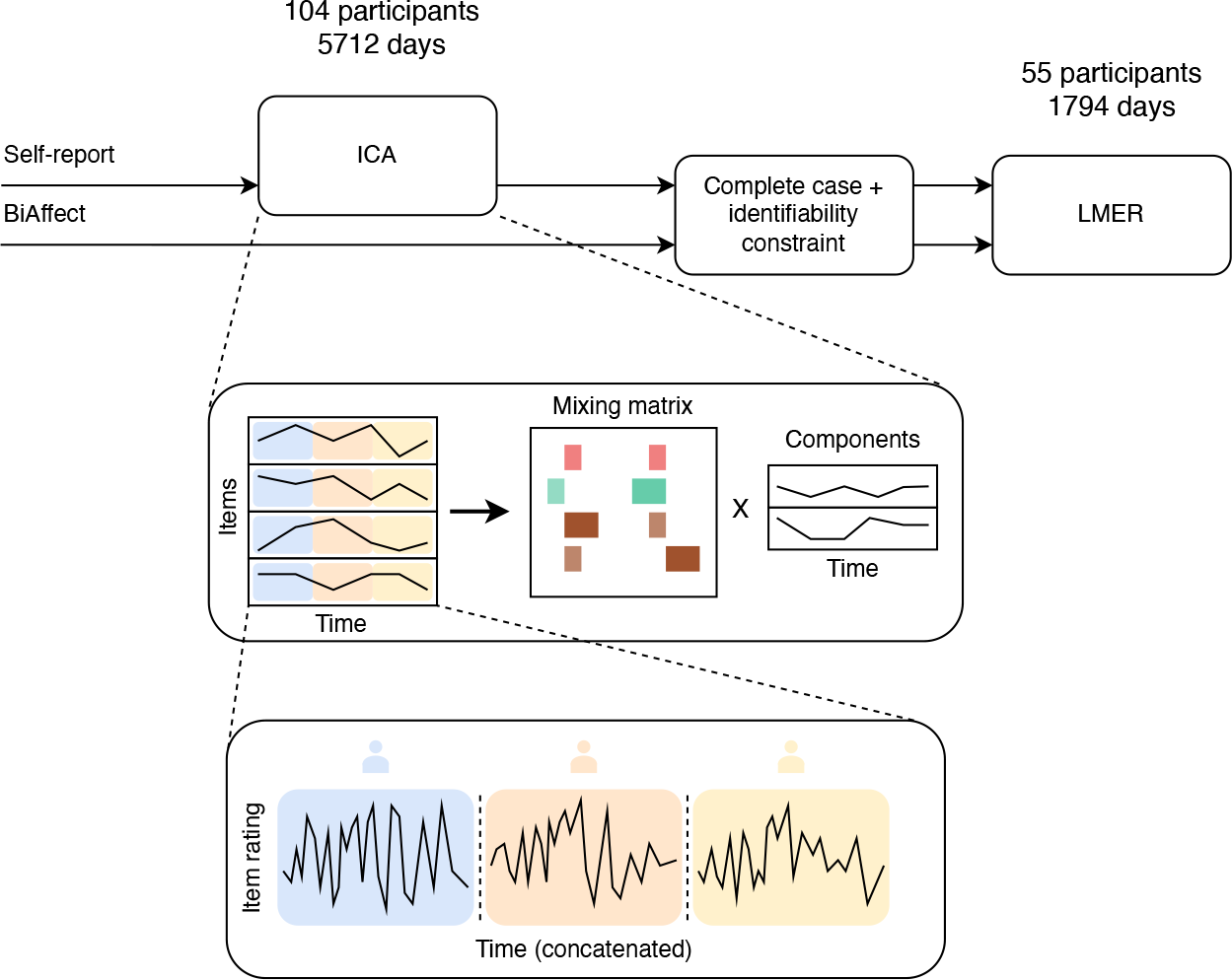
Overview of analysis pipeline. The ICA diagram displays how a collection of multiple time series (one series for every self-report item) gets decomposed into a mixing matrix (depicted as a bar diagram) and a reduced number of independent components with the time domain kept intact. Every item time series is constructed by concatenating the individual participant time series. The complete case and identifiability constraints are specified in the Results. ICA = independent component analysis; LMER = linear mixed-effects regression.

We demonstrate that our method yields a set of interpretable components of self-report data as well as stable associations between these components and keyboard-derived measures in a clinical sample with suicidal ideation.

## Results

### Demographics

Our release of the CLEAR-3 data set contained 109 participants. Missing data patterns are given in Figure 2. Some participants did not have self-report data in their baseline period, which meant that the ICA was run on 104 participants. Their demographics are given in Table 1. For the models, we included all BiAffect data that fell within the range of included self-report data (see Figure 2).

**Table 1:** Participant demographics. The LMER group is a subgroup of the ICA group. ICA = independent component analysis; LMER = linear mixed-effects regression; SD = standard deviation.

|  | ICA | LMER |
| --- | --- | --- |
| N | 104 | 55 |
| Age (mean (SD)) | 25.66 (4.63) | 26.73 (4.87) |
| Race (%) |  |  |
| Caucasian | 49 (47.1) | 26 (47.3) |
| African American | 14 (13.5) | 7 (12.7) |
| Asian | 10 (9.6) | 5 (9.1) |
| Don't Know or More than one race | 28 (26.9) | 14 (25.5) |
| Unknown | 3 (2.9) | 3 (5.5) |
| Ethnicity (%) |  |  |
| Hispanic | 29 (27.9) | 15 (27.3) |
| Non-Hispanic | 73 (70.2) | 38 (69.1) |
| Unknown | 2 (1.9) | 2 (3.6) |
| Education (%) |  |  |
| High school degree, GED, or trade school | 10 (9.6) | 4 (7.3) |
| Post-graduate work | 23 (22.1) | 15 (27.3) |
| Some college or 2-year degree | 28 (26.9) | 13 (23.6) |
| 4-year college degree | 39 (37.5) | 20 (36.4) |
| Unknown | 4 (3.8) | 3 (5.5) |
| Income (%) |  |  |
| <\$15,000 | 12 (11.5) | 6 (10.9) |
| \$15,000-\$34,999 | 17 (16.3) | 10 (18.2) |
| \$35,000-\$79,999 | 39 (37.5) | 19 (34.5) |
| \$80,000-\$100,000 | 12 (11.5) | 5 (9.1) |
| >\$100,000 | 17 (16.3) | 11 (20.0) |
| Unknown | 7 (6.7) | 4 (7.3) |
| Baseline Clinical Categories (%) |  |  |
| Any current depressive disorder | 64 (61.5) | 32 (58.2) |
| Any current anxiety disorder | 63 (60.6) | 30 (54.5) |
| Any current obsessive-compulsive disorder | 11 (10.6) | 4 (7.3) |
| Any current substance use disorder | 17 (16.3) | 7 (12.7) |
| Any current eating disorder | 12 (11.5) | 6 (10.9) |
| Any current trauma-related disorder | 25 (24.0) | 9 (16.4) |

**Figure 2:**
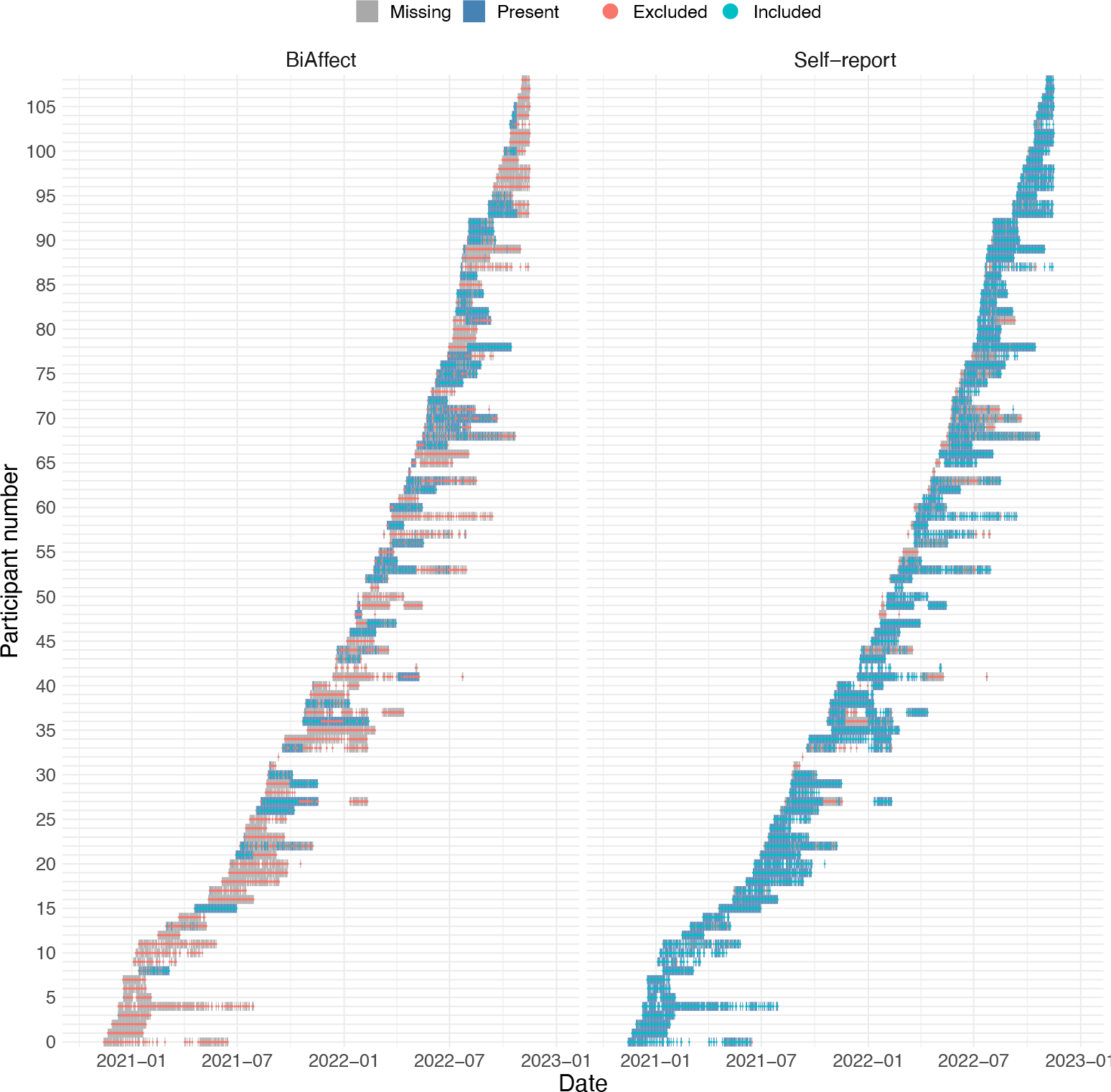
Missingness patterns of BiAffect and self-report data for all participants in the sample. Blank spaces (no grey or blue blocks) indicate that no records whatsoever were available for that date. For the self-report panel, all blocks that are present are also marked as included for the ICA analysis (cyan strikethrough), while those that do not are marked as excluded (red strikethrough). For the BiAffect panel, we included those data in the regression that were both present and fell within the included self-report range.

Linear mixed-effects models require their cases to be complete, i.e., for one day, both BiAffect and self-report features needed to be present. We had a substantial number of incomplete days due to missing keyboard data because participants would sometimes choose to replace the BiAffect keyboard with their own keyboard due to, for instance, multilingual requirements currently not supported by BiAffect. Overall, these incomplete cases resulted in the exclusion of 44 participants. We further required at least two observations for every week within a participant to allow identifiability of the random interaction between week and participant. We therefore excluded an additional 5 participants, leaving us with 55 participants. The demographics of this subgroup are given in Table 1. In total, 5712 days’ worth of data were fed into the ICA, while the mixed-effects models were built with 1794 days.

### Independent component analysis

The mixing matrix for a 5-component temporal ICA solution is shown in Figure 3 (10- and 20-component solutions are shown in Supplementary Fig. 1 and 3). The values of this matrix indicate how much every estimated independent component contributes to the measured values of a self-report item (loading). We selected 34 self-report items from the CLEAR-3 trial that pertained to various aspects of affective, cognitive, and behavioural functioning and were potentially relevant to acute suicidal ideation. These items are subsets of the Daily Record of Severity of Problems (DRSP),^16^ Brief Agitation Measure (BAM),^17^ Brief Irritability Test (BITe),^18^ Adult Suicidal Ideation Questionnaire (ASIQ),^19^ Positive and Negative Affect Schedule (PANAS),^20^ Interpersonal Needs Questionnaire (INQ),^21^ and some EMA items derived from a prior study (denoted here as ‘Miscellaneous’ or ‘Misc’). The exact questions corresponding to the self-report items are given in Supplementary Table 1. Since every column in Figure 3 is linked to an independent component, we will refer to them by their component number.

**Figure 3:**
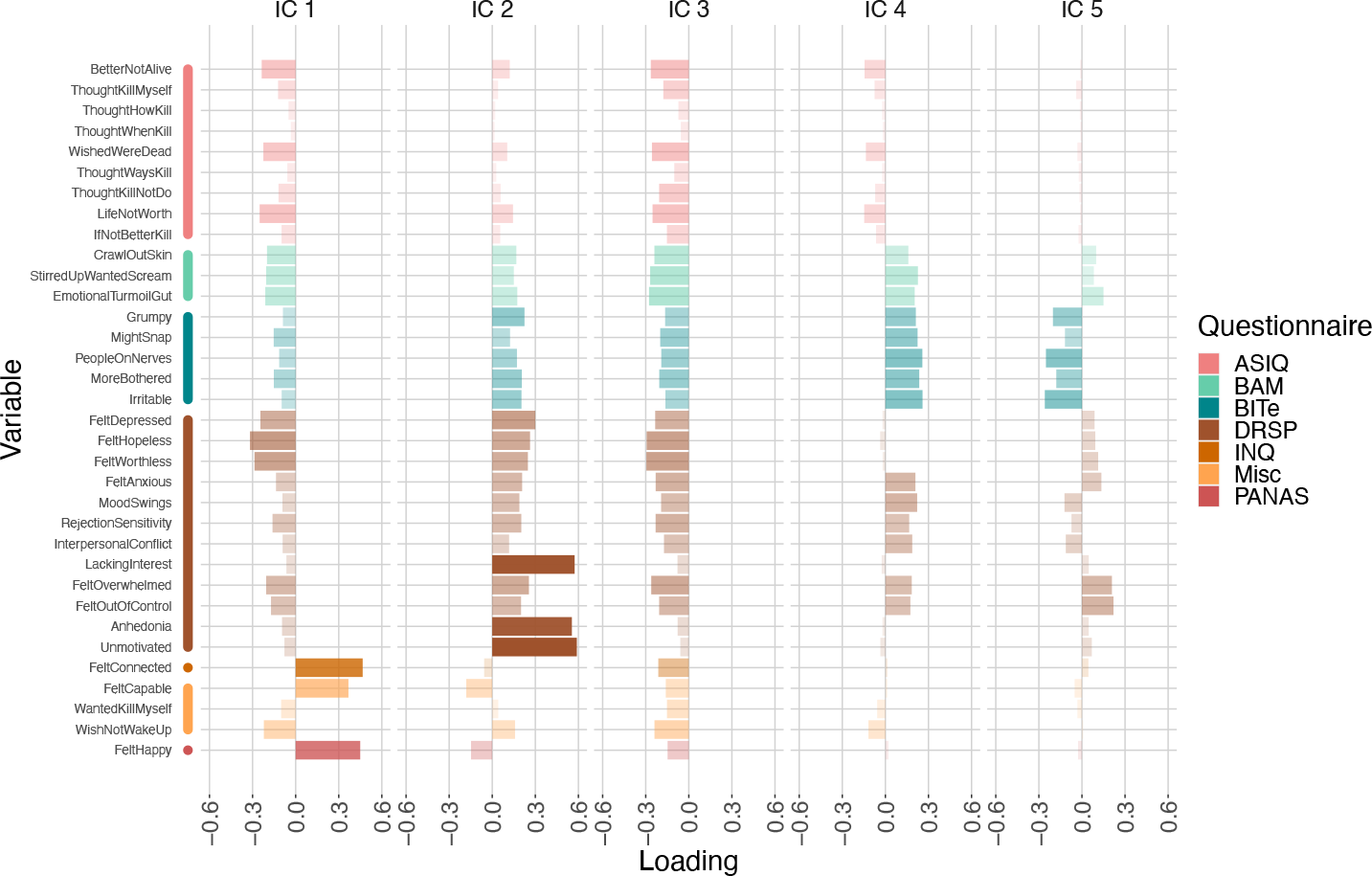
Mixing matrix of the 5-component decomposition of the self-report data. Bar opacity is an additional representation of the loading values. For questionnaire abbreviations, please refer to the main text. IC = independent component.

IC 1 has large positive loadings for the FeltHappy, FeltCapable, and FeltConnected items, which are the only items in our set that pertain to positive affect. The loadings for the rest of the items are in the opposite, negative direction. We will therefore refer to this as the “well-being” component. This polarity pattern reappears for all investigated model orders, up to a reversal of the polarity (Supplementary Fig. 1 and 3). IC 2 shows the same (reversed) pattern, but also displays large loadings for LackingInterest, Unmotivated, and Anhedonia, while the loadings for all other items are comparatively small. We will refer to this IC as the “anhedonia” component. IC 3 shows negative associations with all items in our set, possibly indicating a mean offset of which the intensity varies over time. E.g., if a participant gives consistently lower ratings than other participants, this might be represented with a higher IC 3 intensity. IC 4 gives positive loadings for items measuring agitation (BAM) and the related construct of irritability (BITe), as well as several DRSP items focused on interpersonal reactivity and conflict. We will refer to this IC as the “irritability and social dysfunction” component. Finally, IC 5 displays negative associations with the BITe and small, mixed loadings on the DRSP items. This mix makes it challenging to interpret this component, so we will refrain from naming it.

### Fusion with keyboard dynamics

The BiAffect preprocessing pipeline was based on previous studies.^8,14^ In brief, all keyboard and accelerometery data were aggregated to the daily level. We extracted the following features: 1) median inter-key delay (IKD), an inverse measure of typing speed, 2) 95^th^ percentile IKD, a measure of pausing within typing sessions, 3) mean absolute deviation (MAD) IKD, which quantifies typing speed variability,^8^ 4) autocorrect rate, 5) backspace rate, 6) the total number of key presses per day, 7) the percentage of typing sessions spent upright, and 8) the percentage of typing sessions where the phone recorded movement.

Our mixed-effects models contained fixed effects for all BiAffect features, random intercepts for participants, and random interactions between week and participant. We found no gross violations of model assumptions. For the remainder of this section, we have declared any effects with (corrected) p-values beneath α = 0.05 significant. Forwards-fitting of the random effects indicated that the interaction of week and participant was a significant addition to all models (for all models, p < 0.0001). Model parameter estimates are given in Table 2. After Bonferroni correction, we found that less phone movement corresponded to more anhedonia (IC 2) on the same day (β = -0.12, p = 0.00030). As for terms with p < 0.05 only in the uncorrected case, we found that increased movement rate was associated with greater well-being (β = 0.071, uncorrected p = 0.0051) in the IC 1 model, higher median IKD (slower typing) predicted more anhedonia (β = 0.098, uncorrected p = 0.013) in the IC 2 model, lower median IKD (faster typing; β = -0.094, uncorrected p = 0.030) and a higher total number of key presses (β = 0.062, uncorrected p = 0.027) predicted more irritability and social dysfunction in the IC 4 model, and a lower total number of key presses predicted higher IC 5 intensity (β = -0.060, uncorrected p = 0.025).

**Table 2:**
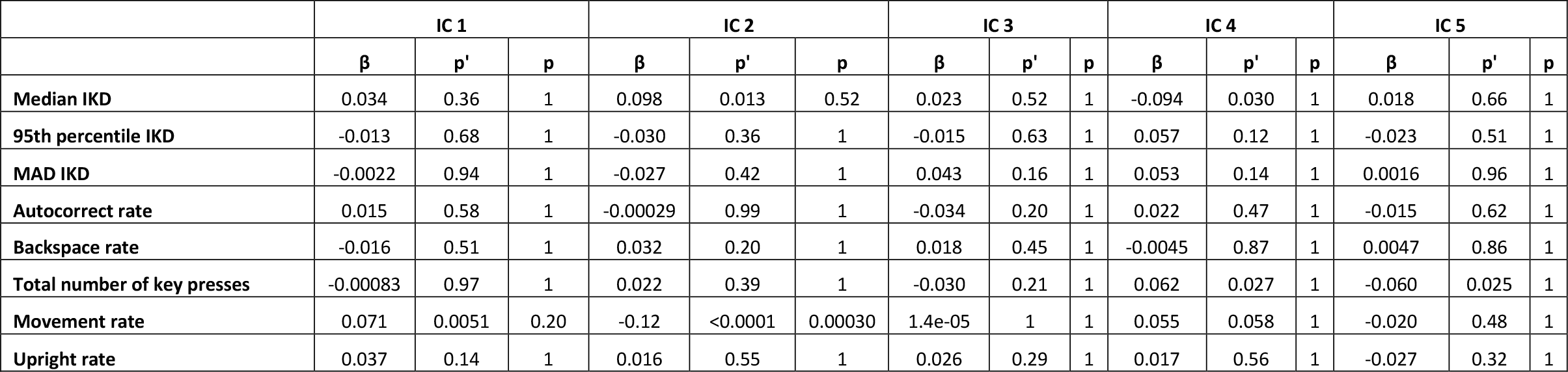
Mixed-effects model estimates with their uncorrected and corrected p values. Every IC corresponds to a separate model. p’ indicates uncorrected p values, p indicates Bonferroni-corrected p values. IC = independent component; IKD = inter-key delay; MAD = mean absolute deviation.

|  | IC 1 |  |  | IC 2 |  |  | IC 3 |  |  | IC 4 |  |  | IC 5 |  |  |
| --- | --- | --- | --- | --- | --- | --- | --- | --- | --- | --- | --- | --- | --- | --- | --- |
| | $\beta$ | $p'$ | $p$ | $\beta$ | $p'$ | $p$ | $\beta$ | $p'$ | $p$ | $\beta$ | $p'$ | $p$ | $\beta$ | $p'$ | $p$ |
| Median IKD | 0.034 | 0.36 | 1 | 0.098 | 0.013 | 0.52 | 0.023 | 0.52 | 1 | -0.094 | 0.030 | 1 | 0.018 | 0.66 | 1 |
| 95th percentile IKD | -0.013 | 0.68 | 1 | -0.030 | 0.36 | 1 | -0.015 | 0.63 | 1 | 0.057 | 0.12 | 1 | -0.023 | 0.51 | 1 |
| MAD IKD | -0.0022 | 0.94 | 1 | -0.027 | 0.42 | 1 | 0.043 | 0.16 | 1 | 0.053 | 0.14 | 1 | 0.0016 | 0.96 | 1 |
| Autocorrect rate | 0.015 | 0.58 | 1 | -0.00029 | 0.99 | 1 | -0.034 | 0.20 | 1 | 0.022 | 0.47 | 1 | -0.015 | 0.62 | 1 |
| Backspace rate | -0.016 | 0.51 | 1 | 0.032 | 0.20 | 1 | 0.018 | 0.45 | 1 | -0.0045 | 0.87 | 1 | 0.0047 | 0.86 | 1 |
| Total number of key presses | -0.00083 | 0.97 | 1 | 0.022 | 0.39 | 1 | -0.030 | 0.21 | 1 | 0.062 | 0.027 | 1 | -0.060 | 0.025 | 1 |
| Movement rate | 0.071 | 0.0051 | 0.20 | -0.12 | <0.0001 | 0.00030 | 1.4e-05 | 1 | 1 | 0.055 | 0.058 | 1 | -0.020 | 0.48 | 1 |
| Upright rate | 0.037 | 0.14 | 1 | 0.016 | 0.55 | 1 | 0.026 | 0.29 | 1 | 0.017 | 0.56 | 1 | -0.027 | 0.32 | 1 |

### Supplementary analyses

We investigated the stability of the ICA solutions across multiple FastICA restarts and found that in most cases the well-being and anhedonia components would combine into one component that indicated general affect (Supplementary Fig. 5). Phone movement remained a significant predictor of anhedonia. When the ICA solutions featured the general affect component instead, we found that phone movement also significantly predicted the general component.

To verify that IC 3 represented a mean offset of self-report responses for certain participants, we correlated the average responses with the IC 3 values and found a negative correlation (Supplementary Fig. 7).

## Discussion

In this work, we introduce a generic method for the analysis and integration of digital phenotyping with self-report data. It utilises temporal ICA to extract interpretable components from the data while keeping the temporal dimension of the data intact, providing a principled method to align different data modalities. We validated the method in a dataset acquired from participants with a history of suicidal ideation and found well-being, anhedonia, and irritability and social dysfunction components in the high-dimensional self-report data. This low-dimensional representation could be predicted by smartphone typing dynamics and accelerometery in that more phone movement while typing was associated with less anhedonia. To our knowledge, this is the first study to demonstrate that passively collected smartphone keyboard dynamics are predictive of a low-arousal state in people with suicidal ideation, as measured using extensive validated instruments that are hard to deploy at scale.

Our method aligned self-report and keyboard dynamics data, allowing their joint analysis and providing further evidence for the use of keyboard dynamics as an ecologically derived marker of mental well-being. Moreover, we distilled high-dimensional data into interpretable components that can be related to the existing literature more easily.

For instance, the emergence of the wellbeing, anhedonia, and irritability and social dysfunction components, as identified by our temporal ICA, can be interpreted in the light of the core affect framework.^22^ According to the core affect theory, core affect is “a neurophysiological state that is consciously accessible as a simple, non-reflective feeling” and is a blend of two dimensions: Pleasure-displeasure and activation-deactivation.^22^ Our components are easily mapped onto this domain. The well-being component aligns mainly with the pleasure dimension and is quite neutral w.r.t. activation. Anhedonia, on the other hand, indicates low activation and a small amount of displeasure. The irritability and social dysfunction component most likely is a blend of high activation and displeasure. Since core affect is postulated to be involved in emotional episodes,^22^ it is encouraging that our temporal ICA is able to identify components that can readily be compared to the core affect dimensions and predicted with data collected passively using smartphones.

Specifically, we showed that phone movement was predictive of the anhedonia component, which is intuitively understandable in that less movement while typing is associated with more anhedonia, a higher lack of interest and a higher lack of motivation. Additionally, we found that phone movement was a trend significant, positive predictor of well-being, which complements the relationship between movement and anhedonia. Since this is the first study investigating the relationship between smartphone keyboard dynamics and suicidality, there is only a small set of prior literature to compare our findings to. Zulueta et al., for example, reported that more phone movement (calculated differently than in our study) predicted higher depression and mania ratings in a sample of participants with bipolar disorder.^6^ However, they also pointed out that bipolar depression can manifest as either psychomotor retardation or agitation,^23^ and therefore their results may not be directly comparable to our findings.

In addition, we found the trend significant effect that larger IKDs (slower typing) predict more anhedonia and that smaller IKDs (faster typing) are related to increased levels of irritability and social dysfunction. While we refrain from drawing conclusions due to the lack of prior literature, it is notable that our findings conform to intuition. Our data also suggest that more key presses predict higher levels of irritability (IC 4 and 5). We note that previous digital phenotyping studies have used measures related to data quantity (e.g., the duration of periods of successful data collection) to detect schizophrenia severity and relapses,^24,25^ which suggests the utility of employing typing dynamics quantity metrics for similar purposes. This would be an interesting direction for future work.

There are several caveats and limitations that we would like to highlight. First, the more components we request from the ICA, the more challenging their interpretation becomes. Many components of the 10- and 20-component solutions, for instance, contain a mix of positive and negative loadings for items from a single scale, which would suggest that such components represent either very specific aspects of a domain or just capture noise (Supplementary Fig. 1 and 3). On the other hand, we stress that low-dimensional ICA solutions need not be the optimal ones. Other decompositions might be equally valid, depending on the level of granularity one wishes to examine.^26^

Second, Figure 2 showed that in some participants substantial portions of the BiAffect data are missing. Indeed, some participants strongly preferred the autocorrect behaviour of the native iOS keyboard, whilst others had multilingual requirements that were not supported by the current version of the BiAffect keyboard. These limits mainly exist because we developed BiAffect and its autocorrect functionalities in-house; they should not be inherent to keyboard typing dynamics itself. We also point out that high proportions of missing data are prevalent in most digital phenotyping studies. With ICA as the core part of our processing pipeline, we can handle this missingness under stationarity conditions. Moreover, the fact that, due to our study incentivisation, the proportions of missingness for data that require active participation (self-report) are much lower than those of passively collected data (BiAffect) is an exception rather than the norm compared to other studies.^25^

Third, while we have found that phone movement while typing is predictive of anhedonia, it is not entirely clear if and how our anhedonia component contributes to suicidal ideation. Links have been found between arousal and suicidal behaviour,^27^ but more research is needed to determine how our anhedonia component maps onto the arousal operationalisations used in the literature. Once this mapping has been clearly delineated, we can potentially leverage the fluctuations in phone movement as part of an early warning system for heightened levels of suicidal ideation.

Finally, the digital phenotyping analysis toolbox is still far from complete. For instance, not much is known about the autocorrelation properties of keyboard and accelerometery dynamics. While our previous research has identified diurnal patterns in keyboard dynamics,^8^ it is not unlikely that there are weekly, monthly, or even seasonal patterns in the BiAffect features, warranting further research.

To conclude, temporal ICA is an effective tool to decompose high-dimensional, daily self-report data without collapsing the time domain. In our dataset containing affective self-report data of people assigned female sex at birth with a history of suicidal ideation, we found ICA-based representations of affect that mapped onto digital phenotyping measures in an interpretable fashion, namely as wellbeing, anhedonia, and irritability and social dysfunction components consistent with the pleasure and activation axes found in core affect theory. We consider this method to be widely applicable and a valuable contribution to the methods toolbox for analysing densely sampled longitudinal and digital phenotyping data.

## Methods

### Study design

Our study utilised data from the CLEAR-3 trial, a randomised controlled crossover trial that investigated how perimenstrual administration of estradiol (E) and progesterone (P), relative to natural steroid withdrawal under placebo, impacts menstrual cycle exacerbation of suicidal ideation and affective symptoms (NCT04112368). The study was approved by the UIC Institutional Review Board. Data acquisition for this study was ongoing, so only baseline (pre-experimental) data were used for this study, which consisted of at least a full menstrual cycle.

### Recruitment and exclusion criteria

All participants were assigned female sex at birth (AFAB), reported past-month suicidal ideation (SI), and were in outpatient treatment. Participants, who were recruited from the community via social media ads and received up to US$1,250 after completing the entire trial, were 18 to 45 years of age, had normal menstrual cycles (25-35 days), did not take any hormonal medications, and had normal weight (BMI 18-29). Exclusion criteria included any long-term nonpsychiatric health condition, a history of hospitalisation for mania or psychosis, or any affective or substance use disorder deemed likely to interfere with safe participation in the clinical trial. All participants provided informed consent for study participation.

### Keypress data preprocessing

The keypress data were aggregated in two steps. First, individual keypresses were aggregated into typing sessions, which begin as soon as the user presses the first key and end when the keyboard is no longer displayed or after six seconds of inactivity.^14^ For each session, the number of autocorrect and backspace presses were counted and divided by the total number of keypresses in the session to get the autocorrect and backspace rates. In addition, the total number of keypresses was counted. Finally, the inter-key delays (IKDs) were calculated between all successive alphanumeric keypresses in the session. From these IKDs, we calculated 1) the median IKD, an inverse measure of typing speed, 2) the 95^th^ percentile IKD, a measure of pausing within sessions, and 3) the mean absolute deviation (MAD) IKD, which quantifies typing speed variability.^8^

After session-level aggregation, the sessions were aggregated to the daily level by taking the mean of all session-level variables. The one exception to this rule were the total numbers of session key presses, which were simply summed across the day. Any days with less than 750 key presses were excluded from further analysis to ensure proper feature estimation.

Finally, the number of key presses was log-transformed and all BiAffect features were standardised w.r.t. the entire sample (i.e., grand mean set to 0 and overall variance to 1) to aid model fitting.

### Accelerometer data preprocessing

Our accelerometer data only included samples collected while the participant was typing on the BiAffect keyboard. While collecting data all throughout the day would yield more data, it would also be more taxing for the smartphone battery, and offloading the accelerometer recording to an external device would require participants to wear an extra device.

Accelerometer data were grouped into typing sessions and low-pass filtered using a second-order bidirectional Butterworth filter with a cutoff frequency of 4 Hz to remove noise.^14^

Afterwards, every sample within a session was classified as either moving (also ‘active’) or stationary based on the magnitude of the filtered x, y, and z accelerometer readings. Magnitudes close to 1 reflect the natural gravitational pull of the earth and therefore indicate the user’s phone is at rest. We classified samples with a magnitude below 0.95 and above as active, and everything that fell within these (inclusive) bounds as stationary. An entire session was classified as active if over 8% of its constituent samples was classified as active. In addition, each session was classified as upright or not using the median values of the filtered x and z values. If the median z value of a session was (strictly) below 0.1 and the median x value was in-between -0.2 and 0.2 (inclusive), the session was classified as upright.^14^ Sessions that were not classified as upright could potentially indicate that participants were using their phone while lying down.

Finally, by counting the number of active and upright sessions within a day and dividing those counts by the total number of sessions within a day, we get a rate of active and a rate of upright sessions per day.

### Independent component analysis

The Supplement gives a mathematical description of how a temporal independent component analysis (ICA) factorises data into a mixing and a source matrix. We used the FastICA algorithm to estimate these matrices from the original self-report data.^10^ More specifically, we used the parallel version with *G* set to the log cosh function and *a*_1_ = 1, implemented in *R* (version 4.2.2) by the *fastICA* package (version 1.2.3).

ICAs are typically run on continuous data that can take negative values, while our self-report data were strictly positive Likert scales. We therefore log-transformed all self-report measures prior to running the ICA.

Solving for independent components typically necessitates a stochastic optimisation and therefore has associated run-to-run variability. We ran a sensitivity analysis to check the extent of this variability (Supplementary Fig. 5).

### Data fusion models

A mathematical formulation of the data fusion models is given in the Supplement. We used the *nlme* package (version 3.1.160) for *R* to create these models. We determined conformity to the linear mixed-effects model assumptions by visual assessment. To assess normality of the residuals and random effects, we examined their QQ plots. Heteroskedasticity was judged by plotting the standardised residuals versus the fitted values. We constructed a separate model for each IC and employed Bonferroni corrections to avoid multiple comparison problems.

## Data Availability

Deidentified participant data will be made available on reasonable request to the principal investigator of the CLEAR-3 trial, T.A. Eisenlohr-Moul, when provided with an appropriate analysis plan.

## Contributors

LK: Conceptualisation; data curation; software; formal analysis; methodology; visualisation; writing – original draft.

AN: Data curation; writing – review & editing.

IEL: Conceptualisation; writing – review & editing. EI: Software; writing – review & editing.

FH: Software; writing – review & editing.

EN: Methodology; writing – review & editing.

TAE-M: Conceptualisation; data curation; funding acquisition; investigation; writing – review & editing.

CFB: Funding acquisition; methodology; supervision; writing – review & editing.

AFM: Conceptualisation; funding acquisition; methodology; supervision; writing – review & editing.

AL: Conceptualisation; funding acquisition; methodology; supervision; writing – review & editing.

## Declaration of interests

CFB is founding director and shareholder of SBG Neuro Ltd. AL is a cofounder of KeyWise AI, previously served on the Medical Board for Buoy Health, and currently serves as a digital psychiatry advisor for Otsuka, USA.

## Code availability

All analysis source code is freely available on GitHub: https://github.com/Valkje/clear3-ica.

## Acknowledgements

This study was funded by the European Research Council (101001118). The CLEAR-3 trial was funded by a grant from the National Institute of Mental Health (RF1MH120843). CFB gratefully acknowledges funding from the Wellcome Trust Collaborative Award in Science 215573/Z/19/Z and the Netherlands Organization for Scientific Research Vici Grant No. 17854 and NWO-CAS Grant No. 012-200-013.

## Notes

### Author Declarations

The Institutional Review Board of the University of Illinois at Chicago gave ethical approval for this work.

