## Supplement for "Smartphone keyboard dynamics predict affect in suicidal ideation"

### Self-report items

| Shorthand | Question | Questionnaire |
|---|---|---|
| BetterNotAlive | I thought it would be better if I was not alive. | Adult Suicidal Ideation Questionnaire |
| ThoughtKillMyself | I thought about killing myself. | Adult Suicidal Ideation Questionnaire |
| ThoughtHowKill | I thought about how I might kill myself. | Adult Suicidal Ideation Questionnaire |
| ThoughtWhenKill | I thought about when I might kill myself. | Adult Suicidal Ideation Questionnaire |
| WishedWereDead | I wished I were dead. | Adult Suicidal Ideation Questionnaire |
| ThoughtWaysKill | I thought about ways people kill themselves. | Adult Suicidal Ideation Questionnaire |
| ThoughtKillNotDo | I thought about killing myself, but would not do it. | Adult Suicidal Ideation Questionnaire |
| LifeNotWorth | I thought that life was not worth living. | Adult Suicidal Ideation Questionnaire |
| IfNotBetterKill | I thought that if things would not get better I would kill myself. | Adult Suicidal Ideation Questionnaire |
| CrawlOutSkin | I wanted to crawl out of my skin. | Brief Agitation Measure |
| StirredUpWantedScream | I felt so stirred up inside I wanted to scream. | Brief Agitation Measure |
| EmotionalTurmoilGut | I felt a lot of emotional turmoil in my gut. | Brief Agitation Measure |
| Grumpy | I was grumpy. | Brief Irritability Test |
| MightSnap | I felt like I might snap. | Brief Irritability Test |
| PeopleOnNerves | Other people got on my nerves. | Brief Irritability Test |
| MoreBothered | Things bothered me more than they normally do. | Brief Irritability Test |
| Irritable | I felt irritable. | Brief Irritability Test |
| FeltDepressed | Felt depressed, down, or blue. | Daily Record of Severity of Problems - Expanded |
| FeltHopeless | Felt hopeless. | Daily Record of Severity of Problems - Expanded |
| FeltWorthless | Felt worthless or guilty. | Daily Record of Severity of Problems - Expanded |
| FeltAnxious | Felt anxious, keyed up, or on edge. | Daily Record of Severity of Problems - Expanded |
| MoodSwings | Had mood swings. | Daily Record of Severity of Problems - Expanded |
| RejectionSensitivity | Was more sensitive to rejection or my feelings were easily hurt. | Daily Record of Severity of Problems - Expanded |
| InterpersonalConflict | Had conflicts or problems with people. | Daily Record of Severity of Problems - Expanded |
| LackingInterest | Had less interest in my usual activities (e.g., work, school, friends, hobbies). | Daily Record of Severity of Problems - Expanded |
| FeltOverwhelmed | Felt overwhelmed, that I couldn't cope. | Daily Record of Severity of Problems - Expanded |
| FeltOutOfControl | Felt out of control. | Daily Record of Severity of Problems - Expanded |
| Anhedonia | Did not enjoy my usual activities. | Daily Record of Severity of Problems - Expanded |
| Unmotivated | Felt unmotivated to do my usual activities (e.g., work, school, friends, hobbies). | Daily Record of Severity of Problems - Expanded |
| FeltConnected | I felt close and connected with other people who are important to me. | Interpersonal Needs Questionnaire |
| FeltCapable | I felt capable in my daily tasks. | Item from prior EMA study (Misc) |
| WantedKillMyself | I wanted to kill myself. | Item from prior EMA study (Misc) |
| WishNotWakeUp | I wished I could go to sleep and never wake up. | Item from prior EMA study (Misc) |
| FeltHappy | I felt happy. | Positive and Negative Affect Schedule |

***Table S1: Self-report item question content.***

### Independent component analysis

In general, an ICA will decompose a data matrix $X$ into a mixing matrix $A$ and a source matrix $S$ such that:[1]

$$X = AS + \epsilon,$$

where $\epsilon$ represents an error term. $X$ is a $p$ by $n$ matrix, where $p$ indicates the number of self-report items and $n$ corresponds to the number of days of data. In our case, we concatenate the data of all participants along the time axis to 1) ensure $n$ is sufficiently large to run the ICA and 2) receive a single set of independent components that applies to all participants. $A$ is a $p$ by $q$ and $S$ is a $q$ by $n$ matrix, where $q$ indicates the number of components we would like the data to be reduced to. Note that if $q = p$, $\epsilon = 0$. In other words, if we request as many components as there are self-report items, there is no error and $X$ is exactly equivalent to $AS$.

### Linear mixed-effects models

Mixed-effects models divide their effects into fixed effects, which are considered the effects that (in theory) apply to the entire population, and random effects, which represent deviations from the fixed effects that are due to the idiosyncrasies of our specific sample.[2] A typical example of random effects is the participant-specific deviation from the global mean in a repeated-measures experiment. In our study, the self-report data did indeed show such participant-specific deviations. Moreover, we found evidence that, depending on the participant, there were week-to-week deviations as well. Such deviations are not unsurprising, as a specific week might have been good for some participants, while others might have experienced it as a particularly bad one. The menstrual cycle of our participants is also likely to bring about periodic fluctuations in the self-report data that can be captured on the weekly level. We therefore opted for modelling a random effect of week nested within participants. In other words, we estimate a random intercept per participant, as well as a random interaction between week and participant. We can formulate a model of $N_{part}$ participants with a participant-dependent $N_{week}$ number of weeks as:

$$y_{ijk} = \beta_0 + b_i + b_{ij} + \boldsymbol{\beta_1^T x_{ijk}} + \epsilon_{ijk},$$

where $i = 1, \ldots, N_{part}$ is the participant index, $j = 1, \ldots, N_{week}$ is the week index, and $k = 1, \ldots, 7$ is the day-of-week index. $y_{ijk}$ represents an independent component (IC) value for participant $i$ in week $j$ on day $k$ and is our dependent variable. $\beta_0$ is the grand mean of the IC values across all participants and weeks. $b_i$ denotes the random effect of participant, i.e., how much participant $i$ shifts the grand mean, on average. Similarly, $b_{ij}$ indicates how much week $j$ shifts the participant-specific mean $\beta_0 + b_i$, but only for participant $i$. $\boldsymbol{x_{ijk}}$ is the BiAffect feature (column) vector, which holds a value for the inter-key delay, autocorrect rate, phone movement rate, et cetera. Like $y_{ijk}$, it is specific to participant $i$ in week $j$ on day $k$. In contrast, the parameter vector $\boldsymbol{\beta_1^T}$ does not depend on a specific participant, week, or day: This vector represents our fixed effects. (The superscripted $T$ denotes the transpose, converting the column vector into a row vector.) Finally, the model has an error term $\epsilon_{ijk}$, which incorporates all IC variation that is not captured by the rest of our model.

We assume that all random effects and the error term are normally distributed around 0. In other words:

$$b_i \sim \mathcal{N}(0,\, \sigma_1^2), \quad b_{ij} \sim \mathcal{N}(0,\, \sigma_2^2), \quad \epsilon_{ijk} \sim \mathcal{N}(0,\, \sigma^2),$$

where $\sigma_1^2$, $\sigma_2^2$, and $\sigma^2$ represent the variance of, respectively, the participant random intercept, the random interaction between participant and week, and the error.

**Model order analysis**

In the main text, we discussed the most parsimonious 5-component solution. Different ICA model orders might be just as valid, however. To examine the behaviour of the independent components across multiple model orders, we reran our analyses with ten and twenty components and correlated the independent components of those solutions with the independent components of the 5-component solution.

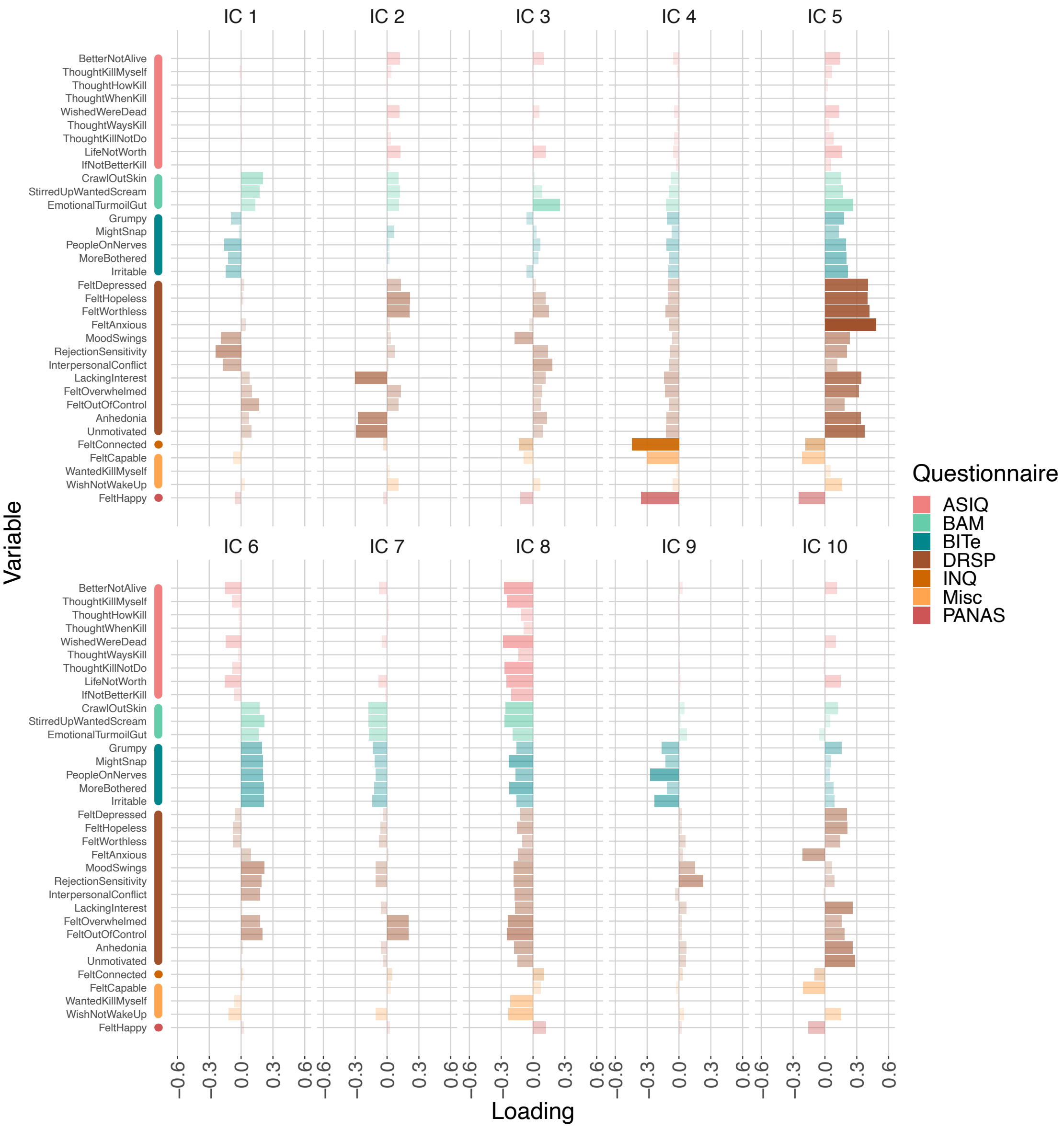

***Figure S1: Mixing matrix of the 10-component ICA solution.*** *IC = independent component. For the questionnaire abbreviations, please refer to Table S1.*

**Ten components**: The mixing matrix of a 10-component ICA solution is given in Figure S1: Mixing matrix of the 10-component ICA solution. IC = independent component. For the questionnaire abbreviations, please refer to *Table S1*. and the cross-correlations between the independent components of this solution and the original 5-component solution are given in Figure S2: Cross-correlations between the independent component time series of the 5- and 10-component solutions. IC = independent component.. To facilitate the comparison between the ICs of these solutions, here we will refer to the components by the prefix $IC_X$, where the subscript X indicates the model order (e.g., $IC_5$ for an IC of the 5-component solution). $IC_5$ 1 correlates most strongly with $IC_{10}$ 4, which is expected given the large loadings on the well-being variables in their respective mixing matrices. (Note that the sign of the correlations does not matter, as the independent components themselves are only defined up to a multiplicative sign.[1]) $IC_5$ 2, on the other hand, shows strongest correlations with both $IC_{10}$ 2 and $IC_{10}$ 5. The correlation with $IC_{10}$ 2 can be explained by its large loadings on the anhedonia items, and the correlation with $IC_{10}$ 5 can be explained by a comparable loading polarity pattern (i.e., negative loadings on positive variables, positive

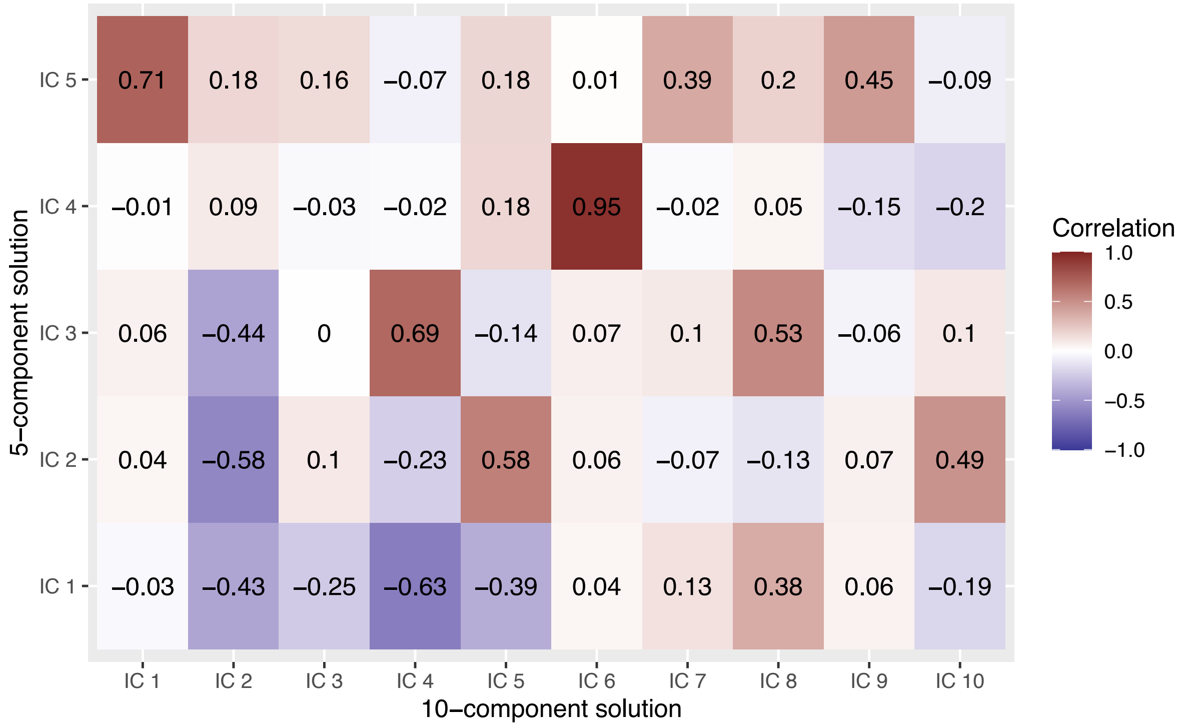

***Figure S2: Cross-correlations between the independent component time series of the 5- and 10-component solutions.*** *IC = independent component.*

loadings on negative variables, and relatively large loadings on the DRSP variables). $IC_5$ 3 shows the strongest correlation with $IC_{10}$ 4, but also a moderate correlation with $IC_{10}$ 8. We see the same pattern as with $IC_5$ 2: $IC_{10}$ 8 has loadings for which the size corresponds to the loadings of $IC_5$ 3, while $IC_{10}$ 4 displays a loading polarity pattern similar to that of $IC_5$ 3. It is not unlikely that $IC_5$ 3 has split up into these two $IC_{10}$ components. $IC_5$ 4 shows a strong correlation with $IC_{10}$ 6, which is not surprising given their similar loading values. $IC_5$ 5, finally, shows its strongest correlation with $IC_{10}$ 1. Their loadings are somewhat comparable, with positive loadings on the agitation items (BAM), negative loadings on the irritability items (BITe), and mixed loadings on the DRSP items.

As for the mixed-effects models of the 10-component solution, none of the effects survived Bonferroni correction (see Table S2). We point out, however, that the nominally significant, negative effect of phone movement on $IC_{10}$ 5 is consistent with what we found for the anhedonia component of the 5-component solution.

All models except the one for $IC_{10}$ 8 showed no signs of heteroskedasticity. Most models, except those for $IC_{10}$ 5 and 10, showed slight departures from normal residuals. Participant-level random effects of the models for $IC_{10}$ 1 and 6-10 displayed small to medium deviations from normality. Week-within-participant-level random effects showed small deviations from normality for all models except for the one for $IC_{10}$ 10.

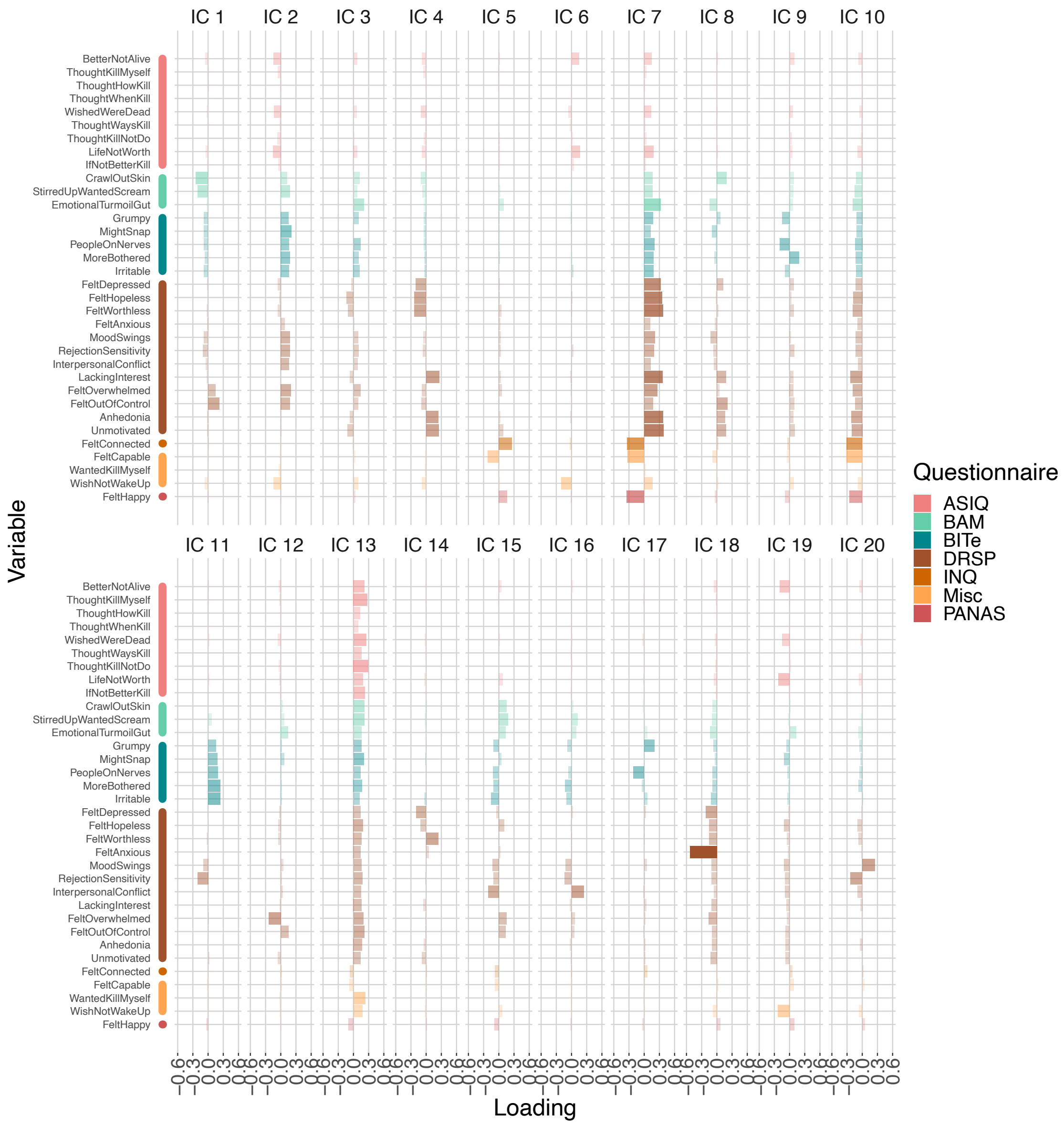

***Figure S3: Mixing matrix of the 20-component solution.*** *IC = independent component. For the questionnaire abbreviations, please refer to Table S1.*

**Twenty components**: The mixing matrix of the 20-component ICA solution is given in Figure S3: Mixing matrix of the 20-component solution. IC = independent component. For the questionnaire abbreviations, please refer to *Table S1*. and the cross-correlations between its components and the components of the 5-component solution are given in Figure S4: Cross-correlations between the independent component time series of the 5- and 20-component solution. IC = independent component.. $IC_5$ 1 and $IC_5$ 2 correlate most strongly with $IC_{20}$ 7. The 20-component mixing matrix shows that $IC_{20}$ 7 has relatively strong loadings on both the well-being and anhedonia items, suggesting that it is a recombination of $IC_5$ 1 and $IC_5$ 2. Our phone movement sensitivity analysis (described below) shows results consistent with this suggestion. $IC_5$ 3 shows its strongest correlations with $IC_{20}$ 10 (similar loading pattern) and $IC_{20}$ 13 (similar loading magnitude), which is the same behaviour we found for the 10-component solution. $IC_5$ 4 has a strong correlation with $IC_{20}$ 2, which is (again) unsurprising because they display very similar loadings. $IC_5$ 5 displays its strongest correlation for $IC_{20}$ 15: Both components again show positive agitation loadings, (mostly) negative irritability loadings, and mixed DRSP loadings.

Consistent with what we found in our sensitivity analysis, the mixed-effects model of the general affect component of the 20-component solution ($IC_{20}$ 7) shows a significant association with phone movement after Bonferroni correction ($\beta = -0.14$, $p = 0.00011$). The sign of this effect is also consistent with the effect of phone movement on the anhedonia component in the 5-component solution ($IC_5$ 2): More movement predicts lower ratings on LackingInterest, Anhedonia, and Unmotivated.

Only the model of $IC_{20}$ 13 showed signs of heteroskedasticity, but several other models ($IC_{20}$ 1, 6, 18, and 20) displayed structure in their residuals versus fitted-values plots. All models except the ones for $IC_{20}$ 3, 7, and 18 showed small to medium deviations from residual normality. Participant-level random effects deviated moderately from normality for all models except the ones for $IC_{20}$ 9, 10, 11, and 18. Week-within-participant-level random effects deviated slightly from normality for all models except the ones for $IC_{20}$ 7, 18, and 20.

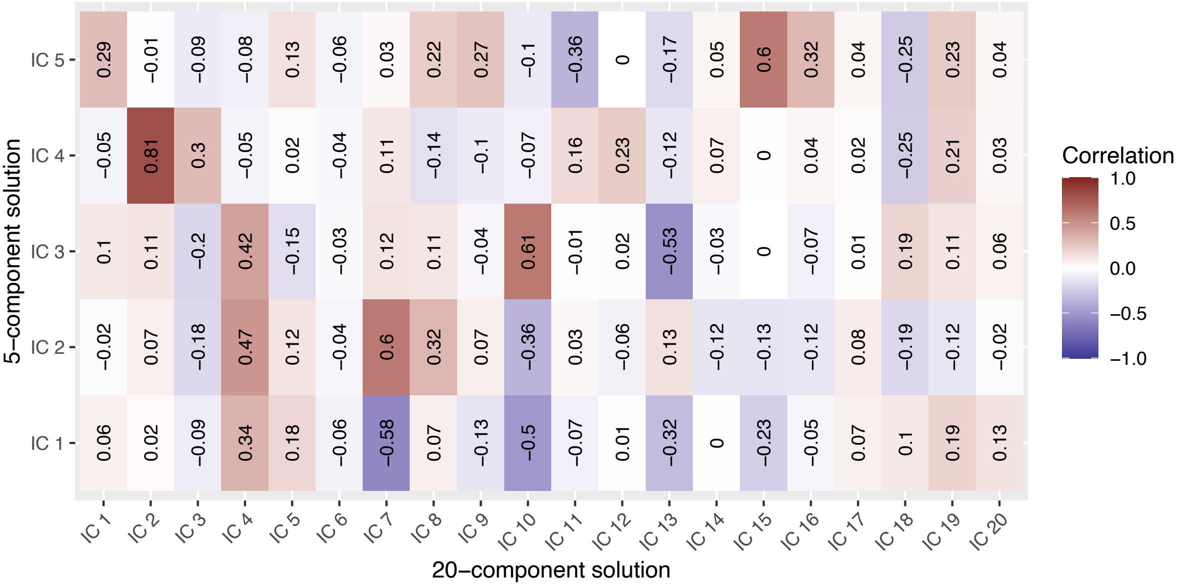

***Figure S4: Cross-correlations between the independent component time series of the 5- and 20-component solution.*** *IC = independent component.*

| | IC 1 | | | IC 2 | | | IC 3 | | | IC 4 | | | IC 5 | | | IC 6 | | |
|---|---|---|---|---|---|---|---|---|---|---|---|---|---|---|---|---|---|---|
| | **β** | **p'** | **p** | **β** | **p'** | **p** | **β** | **p'** | **p** | **β** | **p'** | **p** | **β** | **p'** | **p** | **β** | **p'** | **p** |
| **Median IKD** | 0.0051 | 0.91 | 1 | -0.11 | 0.0091 | 0.73 | -0.10 | 0.029 | 1 | 0.012 | 0.74 | 1 | 0.070 | 0.076 | 1 | -0.088 | 0.040 | 1 |
| **95th percentile IKD** | -0.043 | 0.25 | 1 | 0.046 | 0.20 | 1 | -0.00049 | 0.99 | 1 | -0.0049 | 0.87 | 1 | -0.0067 | 0.84 | 1 | 0.053 | 0.14 | 1 |
| **MAD IKD** | -0.00060 | 0.99 | 1 | 0.018 | 0.61 | 1 | 0.11 | 0.0076 | 0.61 | 0.011 | 0.72 | 1 | -0.056 | 0.092 | 1 | 0.066 | 0.065 | 1 |
| **Autocorrect rate** | 0.0075 | 0.81 | 1 | -0.0095 | 0.76 | 1 | 0.0090 | 0.79 | 1 | -0.033 | 0.20 | 1 | -0.0094 | 0.74 | 1 | 0.018 | 0.56 | 1 |
| **Backspace rate** | 0.018 | 0.53 | 1 | -0.038 | 0.16 | 1 | 0.034 | 0.26 | 1 | 0.011 | 0.62 | 1 | 0.031 | 0.23 | 1 | -0.012 | 0.66 | 1 |
| **Total number of key presses** | -0.057 | 0.048 | 1 | -0.0086 | 0.75 | 1 | -0.056 | 0.060 | 1 | -0.011 | 0.64 | 1 | 0.028 | 0.28 | 1 | 0.059 | 0.033 | 1 |
| **Movement rate** | -0.043 | 0.15 | 1 | 0.045 | 0.12 | 1 | -0.042 | 0.18 | 1 | -0.018 | 0.47 | 1 | -0.079 | 0.0027 | 0.21 | 0.044 | 0.13 | 1 |
| **Upright rate** | 0.0035 | 0.90 | 1 | -0.062 | 0.027 | 1 | -0.0042 | 0.89 | 1 | -0.0062 | 0.79 | 1 | 0.013 | 0.63 | 1 | 0.0028 | 0.92 | 1 |

***Table S2: Model estimates for the 10-component ICA solution.*** *Every IC corresponds to a separate model. p' indicates uncorrected p values. IC = independent component; IKD = inter-key delay; MAD = mean absolute deviation.* *(Continues on next page.)*

| IC 7 | | | IC 8 | | | IC 9 | | | IC 10 | | |
|---|---|---|---|---|---|---|---|---|---|---|---|
| **β** | **p'** | **p** | **β** | **p'** | **p** | **β** | **p'** | **p** | **β** | **p'** | **p** |
| 0.061 | 0.16 | 1 | -0.034 | 0.42 | 1 | 0.054 | 0.24 | 1 | 0.024 | 0.56 | 1 |
| 0.040 | 0.27 | 1 | -0.0044 | 0.90 | 1 | -0.029 | 0.46 | 1 | 0.0024 | 0.94 | 1 |
| -0.037 | 0.30 | 1 | 0.054 | 0.13 | 1 | -0.020 | 0.61 | 1 | 0.012 | 0.74 | 1 |
| -0.025 | 0.43 | 1 | -0.027 | 0.37 | 1 | -0.018 | 0.57 | 1 | -0.026 | 0.38 | 1 |
| -0.0045 | 0.87 | 1 | -0.0058 | 0.83 | 1 | -0.013 | 0.66 | 1 | -0.012 | 0.67 | 1 |
| -0.018 | 0.52 | 1 | -0.040 | 0.14 | 1 | 0.0038 | 0.90 | 1 | -0.0076 | 0.78 | 1 |
| 0.046 | 0.11 | 1 | 0.041 | 0.15 | 1 | -0.023 | 0.45 | 1 | -0.084 | 0.0026 | 0.21 |
| -0.085 | 0.0030 | 0.24 | 0.034 | 0.23 | 1 | -0.0043 | 0.89 | 1 | -0.061 | 0.028 | 1 |

***Table S2 (continued).***

| | IC 1 | | | IC 2 | | | IC 3 | | | IC 4 | | | IC 5 | | | IC 6 | | |
|---|---|---|---|---|---|---|---|---|---|---|---|---|---|---|---|---|---|---|
| | **β** | **p'** | **p** | **β** | **p'** | **p** | **β** | **p'** | **p** | **β** | **p'** | **p** | **β** | **p'** | **p** | **β** | **p'** | **p** |
| **Median IKD** | 0.062 | 0.16 | 1 | -0.074 | 0.10 | 1 | -0.036 | 0.42 | 1 | 0.074 | 0.087 | 1 | 0.027 | 0.55 | 1 | -0.077 | 0.096 | 1 |
| **95th percentile IKD** | 0.035 | 0.34 | 1 | 0.089 | 0.018 | 1 | -0.047 | 0.21 | 1 | -0.049 | 0.17 | 1 | 0.051 | 0.18 | 1 | 0.047 | 0.23 | 1 |
| **MAD IKD** | -0.042 | 0.26 | 1 | 0.044 | 0.24 | 1 | 0.027 | 0.48 | 1 | -0.0085 | 0.82 | 1 | -0.043 | 0.26 | 1 | 0.021 | 0.59 | 1 |
| **Autocorrect rate** | -0.062 | 0.052 | 1 | 0.031 | 0.33 | 1 | -0.057 | 0.074 | 1 | -0.0086 | 0.78 | 1 | -0.029 | 0.36 | 1 | -0.0069 | 0.83 | 1 |
| **Backspace rate** | 0.028 | 0.32 | 1 | -0.033 | 0.25 | 1 | 0.049 | 0.085 | 1 | 0.059 | 0.034 | 1 | -0.011 | 0.69 | 1 | -0.045 | 0.13 | 1 |
| **Total number of key presses** | -0.032 | 0.26 | 1 | 0.065 | 0.024 | 1 | 0.018 | 0.52 | 1 | 0.0023 | 0.93 | 1 | 0.011 | 0.70 | 1 | 0.058 | 0.051 | 1 |
| **Movement rate** | 0.015 | 0.62 | 1 | 0.040 | 0.18 | 1 | 0.064 | 0.032 | 1 | -0.047 | 0.10 | 1 | 0.044 | 0.14 | 1 | 0.018 | 0.55 | 1 |
| **Upright rate** | -0.075 | 0.011 | 1 | -0.021 | 0.47 | 1 | -0.0012 | 0.97 | 1 | 0.080 | 0.0046 | 0.74 | -0.055 | 0.067 | 1 | 0.028 | 0.36 | 1 |

***Table S3: Model estimates for the 20-component ICA solution.*** *Every IC corresponds to a separate model. p' indicates uncorrected p values. IC = independent component; IKD = inter-key delay; MAD = mean absolute deviation.* *(Continues on next page.)*

| IC 7 | | | IC 8 | | | IC 9 | | | IC 10 | | | IC 11 | | | IC 12 | | | IC 13 | | |
|---|---|---|---|---|---|---|---|---|---|---|---|---|---|---|---|---|---|---|---|---|
| **β** | **p'** | **p** | **β** | **p'** | **p** | **β** | **p'** | **p** | **β** | **p'** | **p** | **β** | **p'** | **p** | **β** | **p'** | **p** | **β** | **p'** | **p** |
| -0.017 | 0.67 | 1 | 0.13 | 0.0016 | 0.25 | -0.054 | 0.25 | 1 | 0.050 | 0.19 | 1 | -0.074 | 0.12 | 1 | -0.0036 | 0.94 | 1 | 0.086 | 0.042 | 1 |
| -0.018 | 0.59 | 1 | -0.021 | 0.54 | 1 | -0.046 | 0.25 | 1 | 0.012 | 0.72 | 1 | -0.00016 | 1 | 1 | 0.046 | 0.23 | 1 | 0.023 | 0.51 | 1 |
| 0.044 | 0.20 | 1 | -0.049 | 0.17 | 1 | 0.063 | 0.12 | 1 | -0.024 | 0.45 | 1 | 0.037 | 0.37 | 1 | 0.0082 | 0.83 | 1 | -0.095 | 0.0074 | 1 |
| -0.025 | 0.40 | 1 | 0.018 | 0.56 | 1 | 0.032 | 0.32 | 1 | -0.034 | 0.21 | 1 | 0.023 | 0.48 | 1 | 0.031 | 0.33 | 1 | 0.033 | 0.27 | 1 |
| 0.028 | 0.29 | 1 | -0.019 | 0.49 | 1 | 0.0044 | 0.88 | 1 | -0.0013 | 0.96 | 1 | 0.024 | 0.42 | 1 | -0.0044 | 0.88 | 1 | -0.013 | 0.63 | 1 |
| 0.0073 | 0.78 | 1 | -0.011 | 0.68 | 1 | -0.062 | 0.038 | 1 | -0.0035 | 0.89 | 1 | -0.028 | 0.36 | 1 | 0.012 | 0.69 | 1 | 0.048 | 0.080 | 1 |
| -0.14 | <0.0001 | 0.00011 | 0.012 | 0.66 | 1 | -0.030 | 0.33 | 1 | 0.041 | 0.11 | 1 | -0.0037 | 0.91 | 1 | 0.0074 | 0.81 | 1 | -0.044 | 0.12 | 1 |
| 0.021 | 0.43 | 1 | -0.046 | 0.098 | 1 | -0.067 | 0.029 | 1 | -0.045 | 0.076 | 1 | -0.020 | 0.53 | 1 | 0.019 | 0.52 | 1 | -0.039 | 0.16 | 1 |

***Table S3 (continued).*** *(Continues on next page.)*

| IC 14 | | | IC 15 | | | IC 16 | | | IC 17 | | | IC 18 | | | IC 19 | | | IC 20 | | |
|---|---|---|---|---|---|---|---|---|---|---|---|---|---|---|---|---|---|---|---|---|
| **β** | **p'** | **p** | **β** | **p'** | **p** | **β** | **p'** | **p** | **β** | **p'** | **p** | **β** | **p'** | **p** | **β** | **p'** | **p** | **β** | **p'** | **p** |
| 0.035 | 0.46 | 1 | -0.048 | 0.29 | 1 | -0.099 | 0.043 | 1 | 0.11 | 0.019 | 1 | -0.078 | 0.075 | 1 | 0.022 | 0.62 | 1 | -0.014 | 0.78 | 1 |
| -0.028 | 0.47 | 1 | -0.00094 | 0.98 | 1 | -0.029 | 0.49 | 1 | -0.088 | 0.030 | 1 | 0.0013 | 0.97 | 1 | -0.022 | 0.56 | 1 | -0.022 | 0.59 | 1 |
| -0.025 | 0.53 | 1 | -0.015 | 0.69 | 1 | 0.11 | 0.0065 | 1 | -0.062 | 0.13 | 1 | 0.077 | 0.037 | 1 | 0.012 | 0.75 | 1 | -0.00034 | 0.99 | 1 |
| 0.0048 | 0.88 | 1 | -0.041 | 0.20 | 1 | 0.054 | 0.11 | 1 | -0.057 | 0.086 | 1 | -0.0041 | 0.90 | 1 | 0.0077 | 0.81 | 1 | 0.0010 | 0.98 | 1 |
| 0.016 | 0.58 | 1 | 0.027 | 0.36 | 1 | -0.025 | 0.41 | 1 | 0.019 | 0.51 | 1 | -0.030 | 0.29 | 1 | -0.0013 | 0.96 | 1 | -0.035 | 0.25 | 1 |
| -0.00091 | 0.98 | 1 | -0.041 | 0.16 | 1 | -0.016 | 0.60 | 1 | 0.028 | 0.36 | 1 | -0.023 | 0.41 | 1 | -0.0093 | 0.74 | 1 | 0.046 | 0.14 | 1 |
| 0.018 | 0.57 | 1 | -0.044 | 0.15 | 1 | -0.026 | 0.42 | 1 | -0.036 | 0.25 | 1 | -0.036 | 0.23 | 1 | 0.034 | 0.25 | 1 | -0.041 | 0.20 | 1 |
| 0.084 | 0.0061 | 0.98 | 0.025 | 0.41 | 1 | -0.023 | 0.46 | 1 | -0.029 | 0.35 | 1 | -0.015 | 0.61 | 1 | 0.0065 | 0.83 | 1 | 0.044 | 0.17 | 1 |

***Table S3 (continued).***

**Phone movement sensitivity analysis**

Because the FastICA algorithm starts with a random estimate, its final solution differs from run to run.[1] We conducted a sensitivity analysis to estimate the impact of these differences on the effect of phone movement. More specifically, we ran the *fastICA* function 100 times on our data, manually examined all ICA solutions to select the component that best matched the anhedonia component and constructed a mixed-effects model for every selected component. This procedure resulted in 100 effect estimates of phone movement.

The ICA solutions displayed a dichotomy; an example from each class of solutions is shown in Figure S5: Example mixing matrices of the two classes of ICA solutions. The left panel shows a solution which features the general affect component (IC 1), while the right panel shows the anhedonia component (IC 5). IC = independent component. For the questionnaire abbreviations, please refer to *Table S1*.. 69% of the solutions featured a general affect component, with large loadings on the positive affect variables and large, opposite loadings on the negative

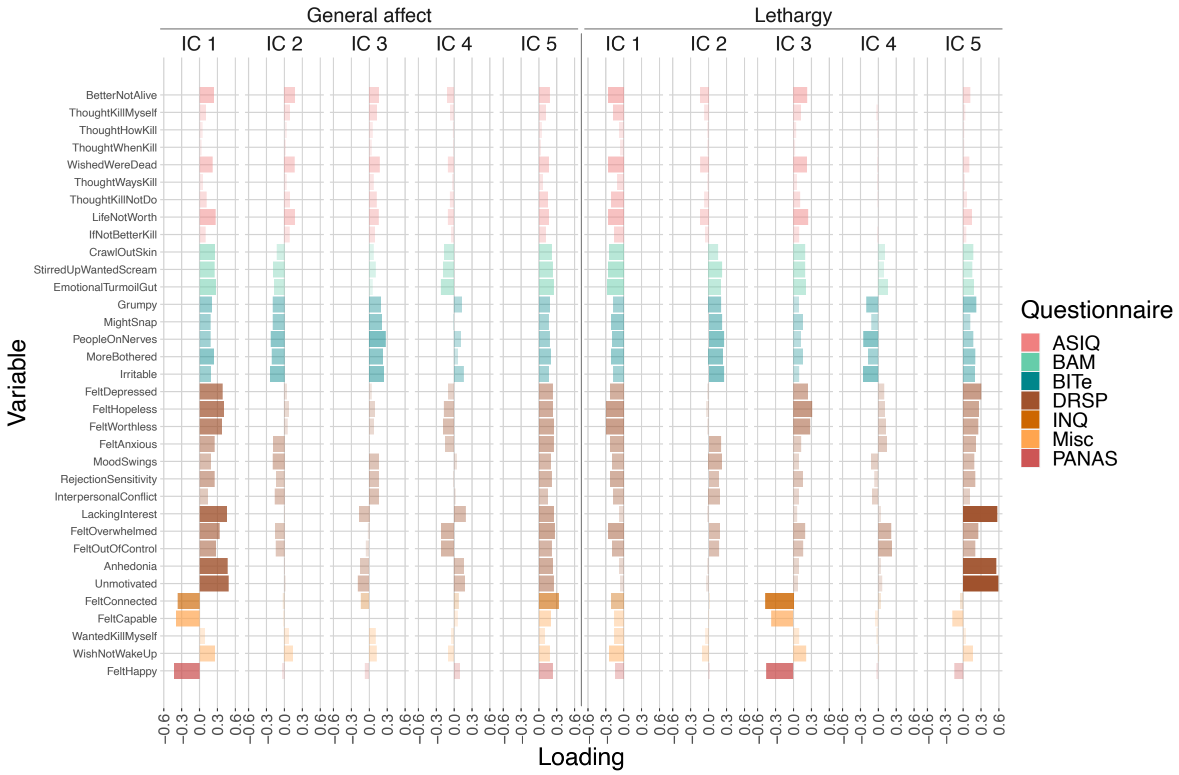

***Figure S5: Example mixing matrices of the two classes of ICA solutions.*** *The left panel shows a solution which features the general affect component (IC 1), while the right panel shows the anhedonia component (IC 5). IC = independent component. For the questionnaire abbreviations, please refer to Table S1.*

affect variables, especially for the DRSP variables. The remaining 31% showed the well-being and anhedonia components previously encountered in the main text, which seem a split of the general affect component.

In the ICA solutions where the anhedonia component did not appear, we created a model of the general component instead. All 100 models reported significant associations with phone movement after Bonferroni correction within a single sensitivity iteration ($-0.14 \leq \beta \leq 0.14$, all $p \leq 0.00031$). The sign of the β values were flipped depending on whether the sign of their associated independent component was also flipped. The size of the β and p values depended on the class of the ICA solution: If the ICA solution contained the general component, $|\beta| = 0.14$ and $p < 0.0001$. Otherwise, $|\beta| = 0.12$ and $p \leq 0.00031$. These results can be interpreted as a dilution of the effect when the general component is split into a well-being and anhedonia one.

Even though the general affect solution was more prevalent in our sensitivity analysis, we still decided to discuss the solution with separate well-being and anhedonia components in our main text because 1) the well-being plus anhedonia solution appeared first in our analyses and 2) the splitting of general affect into well-being and anhedonia provides a more fine-grained view of what drives the association with phone movement.

**Contiguous data analysis**

In the main text, we ran the ICA on all available self-report data. This meant that we also included data from periods with high proportions of missing data, due to which the data stream is not contiguous but fragmented. Temporal ICA is well suited to handle fragmented data since optimising for statistical independence requires assessing the probability density of a source process. ICA does assume, however, that the data were generated from a stationary process. This may not be the case when the distribution underlying data from the periods of fragmentation are very different from the distribution of the contiguous data. Participants might, for example, be less inclined to fill out the self-report items when they are having a bad day and bias the self-report surveys to only measure good days, or vice versa. This non-stationarity could, in turn, affect our ICA results. To test this hypothesis, we reran our sensitivity analysis with the subset of the data that conformed to a contiguity constraint: Self-report data had to be present for at least seven contiguous days. Blocks smaller than seven days would be discarded.

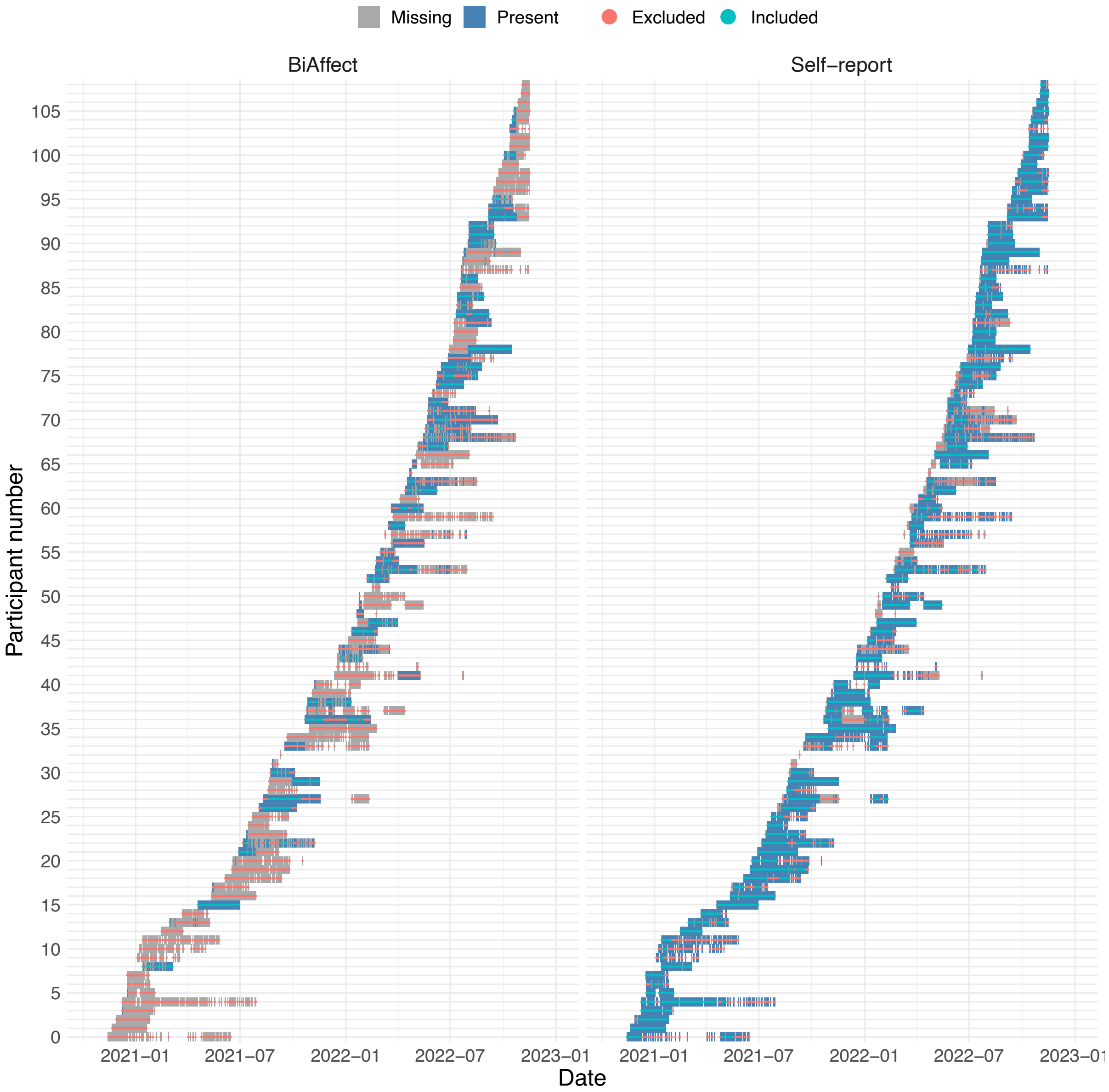

***Figure S6: Missing data patterns with the contiguity constraint.*** *In the right panel, all self-report data that are missing or do not comply with the contiguity constraint are marked as excluded (red strikethrough). In the left panel, all BiAffect data that is missing or falls outside the included range of self-report data is marked as excluded.*

Figure S6: Missing data patterns with the contiguity constraint. In the right panel, all self-report data that are missing or do not comply with the contiguity constraint are marked as excluded (red strikethrough). In the left panel, all BiAffect data that is missing or falls outside the included range of self-report data is marked as excluded. shows what the data inclusion patterns look like when the contiguity constraint is applied. Because some of the self-report data is excluded, some of the BiAffect data also had to be excluded due to the complete-case requirements of the linear regression models. In total, 4215 days' worth of self-report data (98 participants) were fed into the ICA, and 1454 days' worth of BiAffect and ICA component data (47 participants) were fed into the mixed-effects models.

To determine the behaviour of the ICA with contiguous data, we ran the same sensitivity analysis as for the original data. The contiguous ICA solutions showed the same dichotomy as the fragmented ones, but in different proportions: 87% (previously 69%) of the ICA solutions showed the general affect component, while the remaining 13% (previously 31%) showed the split into the well-being and anhedonia components. The solutions with the general component showed a significant effect of phone movement after Bonferroni correction (after rounding, all $|\beta| = 0.12$, all $p \leq 0.00020$); the solutions with the anhedonia component did not ($|\beta| \leq 0.070$, $0.41 \leq p \leq 0.53$). The fact that the proportion of solutions with an anhedonia component has decreased could be interpreted as the consequence of some non-stationarity caused by the fragmented data. Apparently, the fragmented data provide more evidence for separate well-being and anhedonia components, perhaps because the anhedonia items featured more prominently in the fragmented periods. It is, however, also possible that we found the two components more often in the fragmented case simply because that case has more data available to discriminate the two components. The non-significant effects of phone movement on the anhedonia component in the contiguous case contrast with the significant effects in the fragmented case. This contrast can partially be explained by a reduction in statistical power due to the lowered number of data points.

As we pointed out above, if the distributions underlying our data were indeed non-stationary and the periods of missingness in our self-report data were non-random, our entire analysis could be biased towards the days in which participants felt relatively good or bad. Unfortunately, solving the problem of data that are not missing at random (MNAR) is only possible when collecting additional data or making assumptions about the missing data mechanisms.[3] We currently have too little knowledge about what happens to our participants during the periods of missingness and what effect that would have on both the self-report and keyboard dynamics data to properly impute the missing data. For that reason, we decided to do a complete case analysis instead, accepting the potential bias that would introduce.

The model order analysis was also repeated for the contiguous case. For brevity, we do not present the results here, but the patterns in the higher-order ICA solutions are similar to those found in the fragmented case.

**Mean offset component**

In the main text, we found a component with moderate to low negative loadings on all self-report items (IC 3). To examine whether this component might represent a mean offset of self-report ratings for some of our participants, we averaged the self-report ratings per participant and compared them to the component values. More specifically,

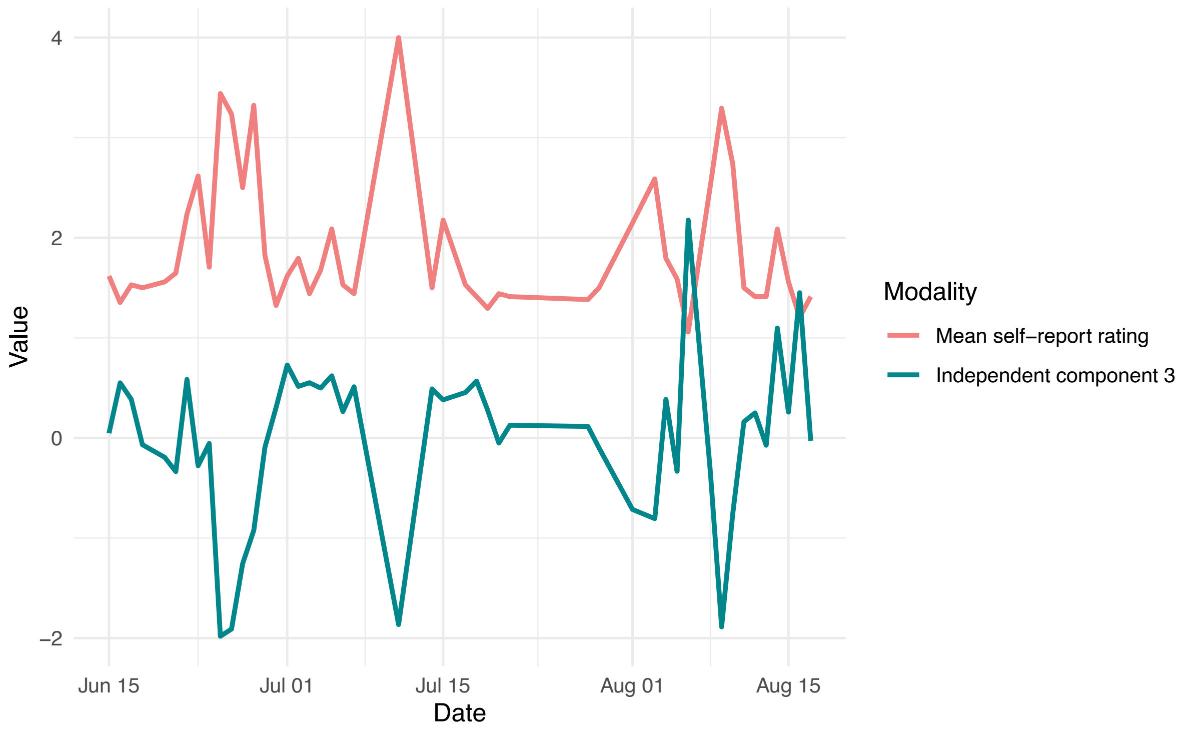

***Figure S7: A participant's average daily rating and IC 3 value over time.***

we first averaged self-report item ratings within participant and within item, and then averaged again within participants but across items. IC 3 values were also averaged within participants. The averaged self-report and component values showed a negative correlation ($r(102) = -0.72$, $p < 0.0001$). For some participants, this relationship was particularly clear (see Figure S7: A participant's average daily rating and IC 3 value over time.). To further examine the effect of the participant means on our ICA solution, we mean-centred all self-report data (including fragmented data) within participants before running the ICA. Figure S8 shows the mixing matrix of the corresponding ICA solution. The component with negative loadings on all items is no longer present, indicating that such a component did indeed represent participant-specific offsets for all self-report items. Moreover, we find further splitting of components found in other ICA solutions. The general affect pattern (negative loadings on negative affect items and positive loadings on well-being items, or vice versa) is still present as IC 3, but the high loadings for the well-being items and anhedonia items have split off into IC 1, 2, and 4, with small to moderate loadings on all other variables. A possible explanation for this behaviour is the fact that the absence of the offset component frees up a component for some additional splitting.

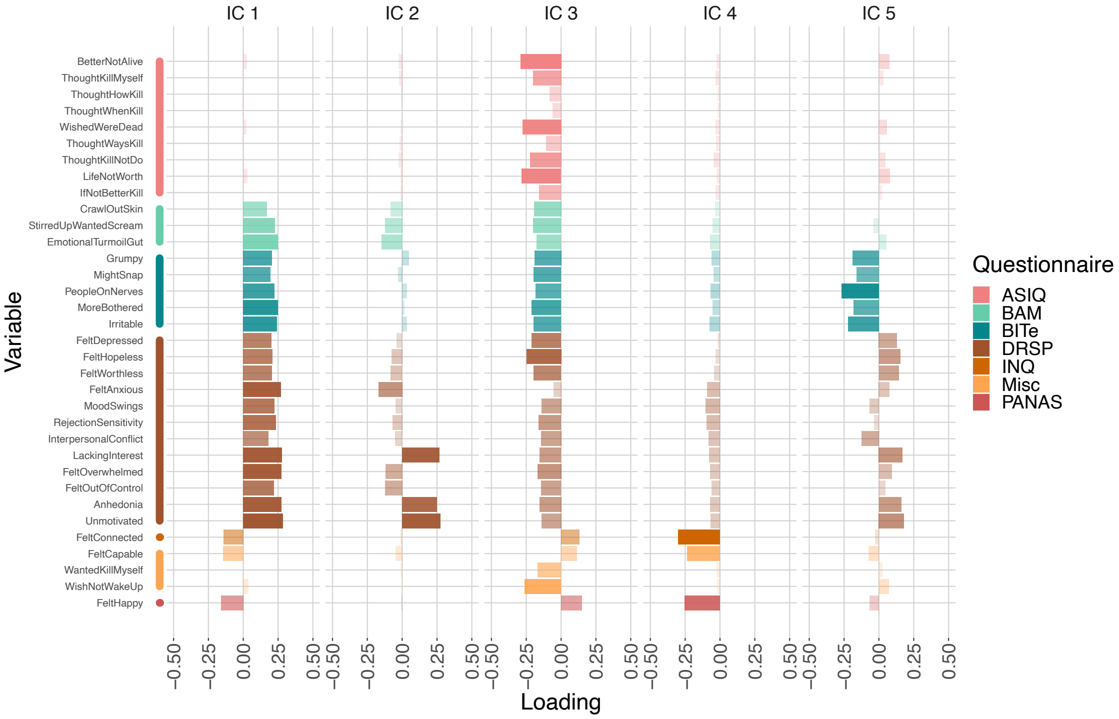

***Figure S8: Mixing matrix of the ICA solution of participant-centred data.*** *IC = independent component.*
